# Lifecourse Socioeconomic Status and Cognitive Performance Among Midlife Latina Women in a California Agricultural Region

**DOI:** 10.1101/2024.05.30.24308219

**Authors:** Jacqueline M. Torres, Lucia Calderon, Katherine Kogut, Kelsey MacCuish, Marcella Warner, Maria T. Rodriguez, Lizari Garcia, Monica Romero, Norma Morga, Aaron K. McDowell-Sanchez, Yesli Perez-Rocha, L. Paloma Rojas-Saunero, Julianna Deardorff, Brenda Eskenazi

## Abstract

A substantial literature evaluates the relationship between lifecourse socio-economic status (SES) and cognitive aging, although most studies primarily focus on non-Hispanic white adults with relatively high SES. Studies that are sufficiently inclusive of minoritized and/or low SES adults typically employ traditional regression approaches, which may not be adequately suited for modeling time-varying lifecourse exposures. We used data from the CHAMACOS Maternal Cognition Study (2022-2024), which included middle-aged, primarily immigrant and Latina women who experienced relatively low lifecourse SES (n = 511). Participants provided information on parental (parental education), childhood (respondent education), and midlife SES (current poverty level), and completed the SOL-INCA neurocognitive assessment, yielded global and domain-specific cognitive performance z-scores. We estimated marginal structural models and evaluated evidence against common lifecourse models (accumulation, critical/sensitive periods, pathways). The overall association between parental education and midlife cognitive scores was attenuated to the null after accounting for respondent education and midlife poverty, in support of the pathways model. Compared to those who completed primary school or less, those who completed any secondary school ({3_*global_cognition*_: 0.19, 95% Confidence Interval [CI]: 0.07, 0.31, 0.32) or high school or above ({3_*global_cognition*_: 0.64, 95% CI: 0.48, 0.80) had higher cognitive z-scores, accounting for both parental education and midlife poverty. The large magnitude of these estimates supports the sensitive periods model. Associations between midlife poverty and cognitive z-scores were modest to null. Our results point to the importance of educational attainment for midlife cognitive health among a subgroup of individuals historically excluded from cognitive aging research.

## INTRODUCTION

There is abundant evidence of a relationship between lifecourse socio-economic status (SES) and cognitive aging.^1-6^ Such associations are evident even in mid-life^2,4,6^, decades before typical dementia onset. However, research that rigorously considers the contribution of SES at varying points across the lifecourse to midlife cognitive outcomes (i.e. appropriately modeling time-varying confounder-mediators)^3,5^ have used data comprised of primarily US-born non-Hispanic White participants. This is notable, given that minoritized racial and ethnic groups in the US face a disproportionate burden of dementia prevalence and incidence.^7^ These prior studies have also focused on the impacts of relatively high levels of life course SES (e.g. high school or college completion). It is not clear whether the midlife cognitive returns to higher SES (e.g. higher education, higher household incomes) are similar for those who have experienced relatively low lifecourse SES.

Research on the impacts of lifecourse exposures – which are, by definition, time-varying -- requires going beyond conventional regression techniques in order to adequately account for the presence of time-varying confounders (see Figure 1).^3,5,8,9^ While there are many studies on lifecourse SES and cognitive aging,^6,10-14^ including studies that center minoritized and/or low SES groups,^12^ estimates from most of these prior studies are either subject to unnecessary residual confounding by omitting measured variables that may serve as common causes of lifecourse SES domains and cognitive outcomes *or* are subject to over-control for potential mediators. Marginal structural models (MSMs) have been well-established in the broader epidemiologic literature as a means of correctly modeling time-varying exposures in service of testing hypothetical lifecourse models^3,5,8,9^ including accumulation, sensitive or critical periods, and pathways models,^15,16^ which are described in greater detail in Table 1. Two our knowledge, only two studies have applied MSMs to the study of lifecourse SES and cognitive aging; again, these studies have been of majority White participants.

**Figure 1.**
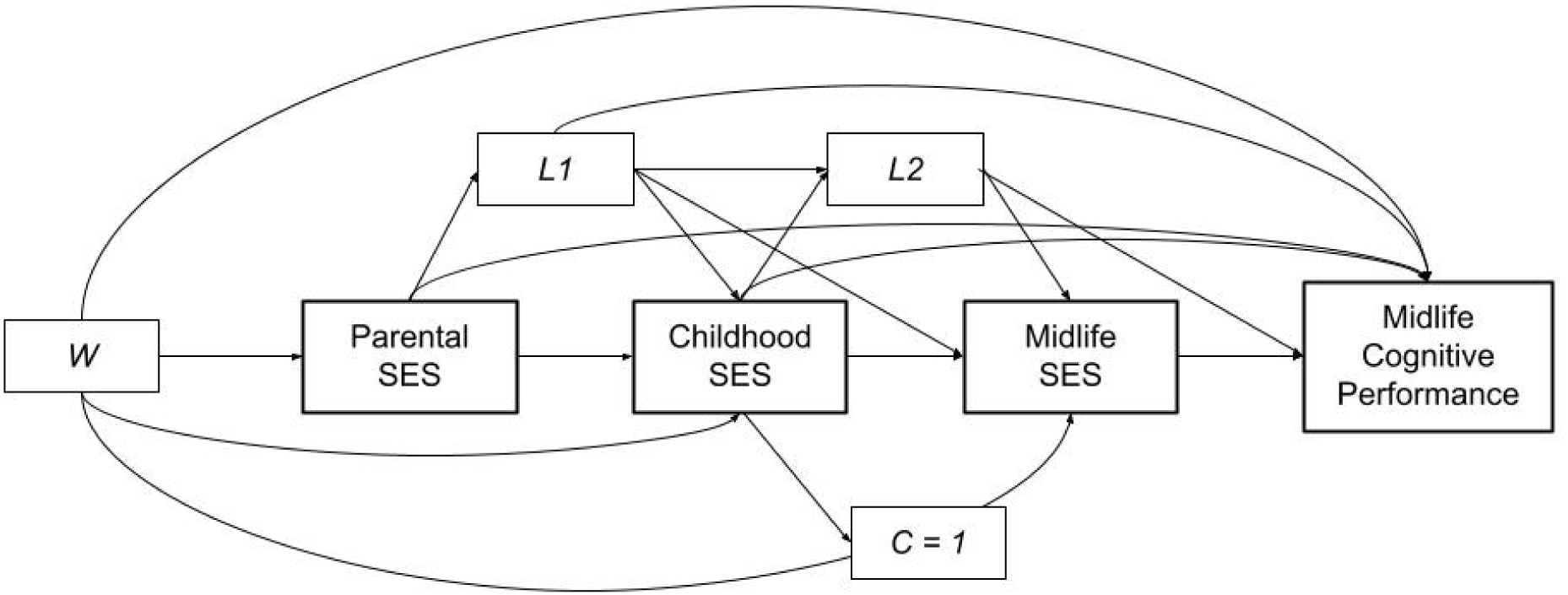
Directed Acyclic Graph depicting hypothetical relationships between socio-economic status at three points along the lifecourse and midlife cognitive performance, along with time-invariant (W) and time-varying (L) confounders, and study attrition between baseline and the 2022/2023 CHAMACOS Maternal Cognition Study visit (C=í). I/V = Age, nativity, language of assessment, *L1* = Adverse childhood experiences, L2 = Marital status, employment, occupational status, chronic health conditions.

**Figure 2.**
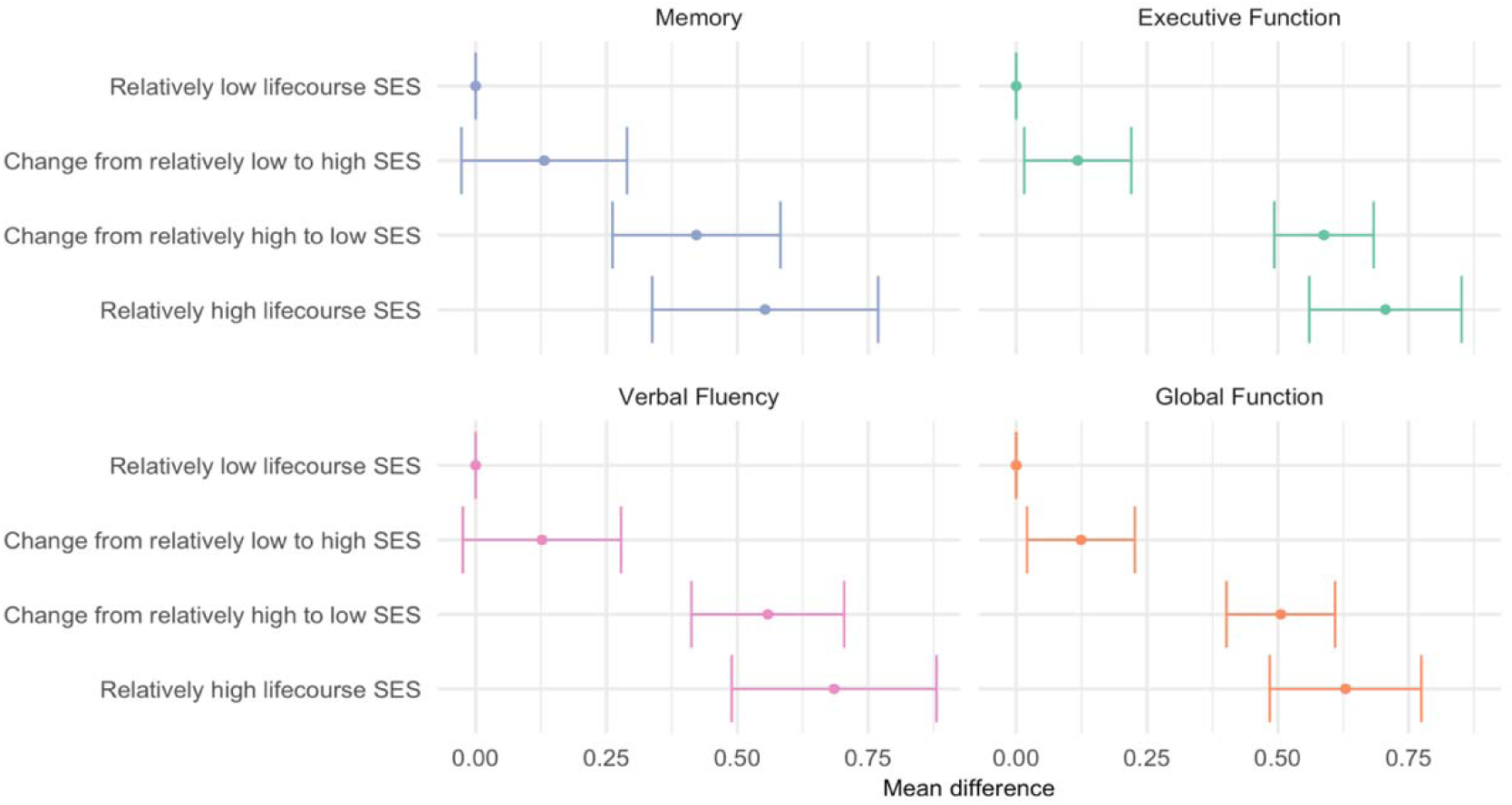
Mean difference in each cognitive outcome between relatively high lifecourse SES, changing from relatively high to relatively low SES across the lifecourse, and changing from relatively low to relatively high SES across the lifecourse, all compared to relatively low lifecourse SES from mid-childhood to midlife (i.e. <=6^th^ grade education and midlife household income at or below the Federal Poverty Level).

**Table 1.** An Overview of Core Lifecourse Models Applied to Relationships between Socio-economic Status and Midlife Cognitive Performance.

|  |  |
| --- | --- |
| Accumulation Model | SES domains at each point across the lifecourse (e.g., from early childhood, mid-childhood, and midlife) have direct effects on later cognitive performance measures (i.e., even after accounting for subsequent lifecourse SES domains). The unique (direct) impacts of each of these SES exposures may accumulate from early to midlife. |
| Sensitive Periods Model | Direct associations between one domain of SES (typically in early or mid-childhood) and cognitive outcomes are of uniquely large magnitude as compared to those observed for associations between other domains of SES and cognitive outcomes. |
| Critical Periods Model | Direct associations are *only* observed for one domain of SES (typically in early or mid-childhood) and not for subsequent measures of lifecourse SES |
| Pathways Model | Only the most recent measure of SES (e.g., in midlife) would have direct effects on midlife cognitive outcomes. There would be no direct influences of SES at earlier time points after accounting for recent SES (e.g. in midlife). |

In the present study, we evaluate the relationship between SES at three points along the lifecourse and cognitive performance outcomes among a sample of midlife and primarily Latina women that have experienced relatively low lifecourse SES (e.g. low levels of intergenerational educational attainment, high levels of poverty). We utilize marginal structural models, estimated via inverse probability weighting, to evaluate core lifecourse hypotheses, i.e., that a) SES in early childhood, mid-childhood, and midlife each have direct associations with midlife cognitive performance (accumulation), b) that direct associations are either present only with early life SES or are of uniquely large magnitude as compared to midlife SES (early critical or sensitive periods), or that c) SES in earlier life only influences midlife cognitive performance via its influence on midlife SES (pathways). In practice, empirical evidence often supports the presence of more than one of these models, which we expect to be the case in our study as well.

## METHODS

Data for this analysis comes from the Centers for the Health of Mothers and Children of Salinas Maternal Cognition Study (CHAMACOS-MCS, 2022-2024), a community-based cohort in California’s Salinas Valley (see Appendix Figure 1 for map) who have been long-time participants in a study focused on environmental exposures and child health.^17,18^ Briefly, in 1999/2000, the study recruited Medicaid-eligible women ≥ 18 years obtaining prenatal care at the Salinas Valley safety net hospital and/or local federally qualified health care centers. Of 1,130 eligible women, 597 enrolled and completed a baseline visit; mother-child dyads were followed thereafter. A refreshment sample of demographically similar mother-child dyads with 9-year-old children was recruited in 2009-2011, culminating in a maternal sample of 635 unique women at that point in time. In 2022-2024, ongoing maternal participants who had completed a study visit between 2014 and 2021 were invited to participate in the CHAMACOS-MCS. Of the 577 participants who met this criterion, 517 completed visits. We excluded 6 respondents missing on socio-demographic covariates; missingness on neurocognitive outcomes varied across measures. Our final analytic sample ranged from n = 495 (for executive function and global cognitive scores) to n = 511 (for memory and verbal fluency scores). A study flow-chart is available in Appendix Figure 2.

We note that CHAMACOS-MCS participants were generally comparable to those who had completed the baseline visits in either 1999/2000 or 2009-2011 but did not participate in the Maternal Cognition Study (Appendix Table 1). Those who remained in the CHAMACOS-MCS were slightly more likely than those who did not to be immigrants, live at or below the federal poverty line, and to have at least one chronic health condition at their baseline visit.

Compared to similarly aged women in the Salinas Valley region, study participants were more likely to have immigrants, to have lower educational attainment, and to be living in poverty (Appendix Table 2). Participants were more comparable to the subset of local middle-aged immigrant women, although were more likely to be living in poverty even compared to this group.

All participants completed written informed consent and study procedures were approved by the University of California, Berkeley Office for the Projection of Human Subjects (Protocol ID 2021-02-14055).

### Measures

#### Lifecourse SES

Parental SES, which is important for shaping the conditions starting in very early life,^19-21^ was measured with an indicator of parental educational attainment. Respondents were asked to report the highest level of education completed by their father and their mother, with the following response options: “none, never attended school”, primary schooling (grades 1-6), junior high (grades 7-9), high school (grades 10-12), high school diploma/GED, some college, college graduate or more, and ‘don’t know/don’t remember’. We created a combined variable indicating the highest level of educational attainment reported across both parents (when there were non-missing values for both parents) or the level of completed educational attainment for the parent with non-missing information (when there were non-missing values for only one parent). Those who reported that they did not know or did not remember the educational attainment of both parents (12% of the analytic sample) were grouped separately, as prior studies have suggested that missing information on parental educational attainment may reflect early-life disadvantage (although the interpretation of the relevant coefficient is not straightforward).^22^

Childhood SES was measured with an indicator of respondents’ educational attainment. Respondents were asked at their baseline visit to report their highest level of educational attainment. In our analytic sample, just under half of respondents (44%) reported receiving a primary education or less (i.e., 6^th^ grade or less). We therefore contrasted this group with those who reported completion of 7^th^ - 11^th^ grade and those who reported completion of high school or GED or higher.

Midlife SES was measured with a binary indicator of whether respondents’ household incomes placed them at or below vs. above the federal poverty level (FPL). Respondents were asked to report their average monthly income over the past 12 months as well as the number of people who were supported by that income. Response options for monthly income were: “$400 or less”; “$401 to 750”; “$751 to 1500”; “$1501 to 2000”, “$2001 to $2500”, and so on, with increasing intervals of $500 up to $5000; “$5001 and above”, or “Don’t know”. We then assigned each participant a monthly income at the mid-point of their responses and compared this to the U.S. Census Bureau’s poverty threshold^23^ for their respective household size to classify individuals as being at or below the FPL vs. above the FPL. A total of 7.2% of the sample were missing income data. For our primary analyses, we created a three-category midlife poverty variable in which those missing income data were assigned their own category. However, in sensitivity analyses, we removed missing observations from our analysis and modeled a binary midlife poverty measure.

#### Neurocognitive Performance Measures

Respondents completed an adapted version of the Study of Latinos – Investigation of Neurocognitive Aging 2 (SOL-INCA2) neurocognitive battery,^24^ which was administered by bilingual and bicultural trained and certified interviewers. See Appendix Text 1 for further details on administration. The assessment included the Brief Spanish English Verbal Learning Test (B-SEVLT),^25,26^ the Digit Symbol Substitution (DSS);^27^ the Trails Making Test (TMT) A and B;^28^ the Digit Span Test Forward and Backward;^29^ the B-SEVLT Delayed Recall; two tasks to measure phonemic (letter) fluency and one task to measure semantic or categorical fluency (see Appendix Table 3 for additional details). Scores on each task was standardized to have a mean of zero and a standard deviation of one (i.e., z-scored) and then summarized into composite z-scores that generally measure domains of memory, executive function, and verbal fluency. We summarized these composite scores into a combined global cognition composite z-score.

A large number (n = 204) of participants were not able to complete TMT B, primarily due to its difficulty^30^; scores were either recoded to the maximum (i.e. worst) possible score or set to missing based on other information (e.g. their reason for discontinuing the task). However, we conducted a sensitivity analysis excluding the TMT B from the calculation of executive function and global composite z-scores.

#### Covariates

We included covariates in our models in a way that aligns with the directed acyclic graph shown in Figure 1. For models of parental educational attainment, we included a measure of respondents’ age in years (non-linear age terms did not improve model fit), a measure that combined information on nativity and age at first migration to the US, and the language they preferred for their neurocognitive assessment (Spanish vs. English). While age at migration is established in adolescence or adulthood for many immigrant women in our sample, it is directly correlated with nativity and therefore incorporated as a combined variable. Given the distributions in the sample we created a 3-category variable of both nativity and age at migration (US-born; immigrant, arrived in childhood or < 18 years; immigrant, arrived in adulthood or ≥ 18 years).

In models of respondent educational attainment, we additionally included a summary measure of retrospectively reported adverse childhood experiences (ACES), coded as 0, 1, 2, 3, 4, or 5+ ACES. In models of midlife (current) poverty, we additionally included indicators of current marital status (married or partnered vs. divorced, separated, widowed, or single), and a binary measure of current work status (i.e. worked outside the home since the last visit). We also included current health status, including a summary measure of the following health conditions (range: 0-5): hypertension, diabetes, heart or blood vessel problems (including heart attack or stroke), cancer, thyroid conditions, and asthma. Hypertension status was based on self-reports of doctor-diagnosed hypertension and in-office measurements of blood pressure (see detailed in Appendix Text 1). Diabetes status was based on self-reports of doctor-diagnosed diabetes as well as fasting glucose and/or either fasting or non-fasting HbA1c measures (see detail in Appendix Text 2). Remaining health conditions were based on self-reports of prior doctor-diagnosis.

#### Statistical Methods

We first calculated summary statistics for the entire analytic sample. We then estimated a series of linear regression models that included main effect terms for each lifecourse SES measure. We adjusted for age, nativity/age at migration, and language of assessment by direct inclusion in the regression model and used inverse probability weights to account for childhood and post-childhood confounders. We generated IPWs from two separate models corresponding to childhood SES (respondent educational attainment) and midlife SES (poverty status). The first model estimated the probability of the participant having above a primary school education (vs. primary school or less) conditional on age, nativity/age-at-migration, language of assessment, parental educational attainment, and the measure of ACEs. The second exposure model estimates two probabilities using ordinal logistic regression: the probability of reporting a household income above the federal poverty level (FPL vs. at or below the FPL) and the probability of missing a household income (vs. at or below the FPL), conditional on all variables included in the first exposure model, respondent educational attainment, and additional socio-demographic and health variables detailed above. We calculated weights from each of these models, each equal to the inverse of the probability of respondents’ true exposure status. These weights were then multiplied together to create a final weight, which we trimmed at the 99^th^ percentile. Models were estimated with robust standard errors.

As described by others,^3^ this model yields estimates of the controlled direct effects of each lifecourse SES measure on midlife cognitive performance. That is, estimates of association between parental educational attainment correspond to those not mediated or explained by respondent educational attainment or midlife poverty. Estimates of association between respondent educational attainment and midlife cognition correspond to those not mediated or explained by midlife poverty and independent of parental educational attainment.

Following prior studies, we additionally estimated the mean differences in cognitive performance scores under the joint lifecourse SES exposures. These were estimated with marginal mean models in which we contrasted cognitive outcomes under hypothetical scenarios of lifecourse SES exposure; 95% confidence intervals were calculated via the bootstrap method. We considered hypothetical scenarios of higher (relative to the rest of the sample) SES across the lifecourse, change from relatively high to relatively low SES, and change from relatively low to relatively high SES, all compared to a scenario of relatively low SES across the lifecourse.

We carried out several sensitivity analyses, including 1) excluding the 6 respondents who completed the neurocognitive assessment by phone, 2) excluding the Trails Making Test B from executive function and global composite scores due, and 3) using a binary measure of poverty status. Finally, we re-estimated models after including inverse probability of attrition weights (IPAWs) to account for attrition from respondents’ baseline visits. These weights were created with models estimating the probability of inclusion in the Maternal Cognition Study conditional on baseline socio-demographic characteristics.

## RESULTS

### Descriptive

Participants in our analysis had a mean age of 48.8 years (SD:5.4) (Table 2). The majority (63.6%) of participants first immigrated to the US during adulthood (41.3% between the ages of 18-24 and 22.3% 25 years or older), with only 10.8% of the participants having been born in the US. Most respondents (88.6%) completed their assessments in Spanish.

**Table 2.** Overall Descriptive Characteristics, CHAMACOS Maternal Cognition Study (n = 511)

|  | n | % or<br>mean<br>(SD) |
| --- | --- | --- |
| **Age, mean (SD)** |  | 48.8 (5.4) |
| **Age of arrival to the US, n (%)** |  |  |
| 0 years | 55 | 10.8 |
| 1-17 years | 131 | 25.6 |
| 18-24 years | 211 | 41.3 |
| 25+ years | 114 | 22.3 |
| **Language of assessment, n (%)** |  |  |
| Spanish | 453 | 88.6 |
| **Highest level of parental educational attainment, n (%)** |  |  |
| No formal educational attainment | 116 | 22.7 |
| Any formal educational attainment | 336 | 65.8 |
| Don't know or don't remember for both parents | 59 | 11.5 |
| **≥ 1 Adverse childhood experience, n (%)** | 277 | 54.2 |
| **Respondent educational attainment, n (%)** |  |  |
| ≤ 6th grade | 224 | 43.8 |
| 7-12th grade | 175 | 34.2 |
| ≥ HS graduate | 112 | 21.9 |
| **Married or living as married, n (%)** | 361 | 70.6 |
| **Worked since last visit, n (%)** | 409 | 80.0 |
| **Poverty categories using 2022 ACS Data, n (%)** |  |  |
| At or below FPL | 182 | 35.6 |
| 100%-200% FPL | 266 | 52.1 |
| >200% FPL | 26 | 5.1 |
| Missing | 37 | 7.2 |
| **Summary health score, n (%)** |  |  |
| No health issues | 189 | 37.0 |
| 1 health issue | 184 | 36.0 |
| 2+ health issues | 138 | 27.0 |

Note: a. The composite health score is based on a count of the following conditions: diabetes, high blood pressure, heart issue, cancer, asthma, and thyroid issue. Diabetes status was based on self-reported history of diagnosis, blood glucose, or HbA1c values; hypertension status was based on self-reported history of diagnosis and in-office blood pressure measurements. The remaining conditions were based on self-reported history of diagnosis. Participants could answer “yes”, “no”, or “don’t know” for each of the self-rated health issues. “Yes” responses were summarized to create the score.

Over half (54.2%) of participants experienced at least one adverse childhood experience. Most (70.6%) women were married or living as married, and approximately 80% had worked since their last study visit. Over one-third of respondents reported no chronic health conditions while just under one-third had 2 or more health conditions.

The majority (65.8%) of participants’ parents had at least some formal educational attainment. Over half of respondents (56.2%) had a 7^th^ grade education or greater (34.2% with 7^th-^ 12^th^ grade education and 21.9% with a high school education or greater); the remaining 43.8% had a 6^th^ grade education or less. Over half of respondents (57.2%) lived above the federal poverty level (FPL), followed by 35.6% of women living at or below the FPL, and 7.2% missing information on poverty status.

#### Associations between Lifecourse SES and Cognitive Outcomes

In models that only included parental educational attainment with adjustment for age, nativity/age-at-migration, and language of assessment (Table 3, Model 1), we observed a positive association between any (vs. no) formal parental educational attainment and respondents’ executive function ({3: 0.33, 95% Confidence Interval [CI]: 0.19, 0.46), verbal fluency ({3: 0.30, 95% CI: 0.12, 0.48), and global cognition ({3: 0.23, 95% CI: 0.10, 0.36) z-scores; associations with memory z-scores were null ({3: 0.05, 95% [CI]: -0.14, 0.24).

**Table 3.** Models of Association between Lifecourse SES and Domain-Specific and Global Cognitive Z-Scores.

| Memory (n = 501) |  |  |  |  |  |  |
| --- | --- | --- | --- | --- | --- | --- |
|  | Model 1 |  | Model 2 |  | Model 3 |  |
|  | Coef | 95% CI | Coef | 95% CI | Coef | 95% CI |
| Parental Education (Ref: none) |  |  |  |  |  |  |
| Any formal education | 0.03 | (-0.16, 0.23) | -0.05 | (-0.24, 0.14) | -0.07 | (-0.25, 0.12) |
| Don't know/ don't remember | -0.21 | (-0.50, 0.07) | -0.19 | (-0.47, 0.09) | -0.26 | (-0.56, 0.03) |
| Respondent Education (Ref: < 6th grade) |  |  |  |  |  |  |
| 7th-11th grade |  |  | 0.34 | (0.17, 0.51) | 0.12 | (-0.05, 0.29) |
| > High School/GED |  |  | 0.40 | (0.20, 0.61) | 0.41 | (0.21, 0.61) |
| Poverty level (Ref: < FPL) |  |  |  |  |  |  |
| > Federal Poverty Level |  |  |  |  | 0.08 | (-0.10, 0.25) |
| Missing |  |  |  |  | 0.11 | (-0.07, 0.30) |
| Executive Function (n = 485) |  |  |  |  |  |  |
|  | Model 1 |  | Model 2 |  | Model 3 |  |
|  | Coef | 95% CI | Coef | 95% CI | Coef | 95% CI |
| Parental education (Ref: none) |  |  |  |  |  |  |
| Any formal education | 0.32 | (0.19, 0.46) | 0.19 | (0.08, 0.31) | 0.18 | (0.06, 0.29) |
| Don't know/ don't remember | 0.12 | (-0.08, 0.32) | 0.13 | (-0.04, 0.31) | 0.10 | (-0.09, 0.29) |
| Respondent education (Ref: < 6th grade) |  |  |  |  |  |  |
| 7th-11th grade |  |  | 0.46 | (0.35, 0.56) | 0.45 | (0.34, 0.56) |
| ≥ High school/GED |  |  | 0.76 | (0.63, 0.89) | 0.81 | (0.68, 0.94) |
| Midlife poverty status (Ref: ≤ FPL) |  |  |  |  |  |  |
| > Federal Poverty Level |  |  |  |  | 0.10 | (-0.02, 0.21) |
| Missing |  |  |  |  | -0.10 | (-0.22, 0.02) |
| Verbal Fluency (n = 501) |  |  |  |  |  |  |
|  | Model 1 |  | Model 2 |  | Model 3 |  |
|  | Coef | 95% CI | Coef | 95% CI | Coef | 95% CI |
| Parental Education (Ref: none) |  |  |  |  |  |  |
| Any formal education | 0.30 | (0.12, 0.49) | 0.18 | (0.01, 0.36) | 0.14 | (-0.03, 0.32) |
| Don't know/ don't remember | -0.03 | (-0.30, 0.24) | -0.08 | (-0.34, 0.19) | -0.05 | (-0.33, 0.24) |
| Respondent education (Ref: < 6th grade) |  |  |  |  |  |  |
| 7th-11th grade |  |  | 0.35 | (0.19, 0.51) | 0.12 | (-0.05, 0.28) |
| ≥ High School/GED |  |  | 0.64 | (0.45, 0.84) | 0.49 | (0.29, 0.68) |
| Midlife poverty status (Ref: ≤ FPL) |  |  |  |  |  |  |
| > Federal Poverty Level |  |  |  |  | -0.07 | (-0.24, 0.11) |
| Missing |  |  |  |  | -0.37 | (-0.55, -0.19) |
| Global Composite (n = 485) |  |  |  |  |  |  |
|  | Model 1 |  | Model 2 |  | Model 3 |  |
|  | Coef | 95% CI | Coef | 95% CI | Coef | 95% CI |
| Parental education (Ref: none) |  |  |  |  |  |  |
| Any formal education | 0.22 | (0.09, 0.36) | 0.12 | (-0.01, 0.24) | 0.08 | (-0.04, 0.20) |
| Don't know/ don't remember | -0.06 | (-0.26, 0.14) | -0.07 | (-0.25, 0.12) | -0.09 | (-0.29, 0.11) |
| Respondent education (Ref: < 6th grade) |  |  |  |  |  |  |
| 7th-11th grade |  |  | 0.36 | (0.25, 0.47) | 0.21 | (0.09, 0.32) |
| > High school, GED |  |  | 0.58 | (0.45, 0.71) | 0.55 | (0.41, 0.68) |
| Midlife poverty status (Ref: ≤ FPL) |  |  |  |  |  |  |
| > Federal Poverty Level |  |  |  |  | 0.03 | (-0.09, 0.15) |
| Missing |  |  |  |  | -0.12 | (-0.25, 0.01) |

Source: CHAMACOS Maternal Cognition Study, 2022-2024. Models of parental educational attainment control for respondent age and nativity/age-at-immigration to the US; models including educational attainment also account for adverse childhood experiences; models that include midlife poverty status also account for current marital status, employment status, depressive symptoms, anxiety symptoms, and chronic health conditions (e.g. hypertension).

In models that also included respondents’ educational attainment (Table 3, Model 2) and midlife poverty status (Table 3, Model 3) and accounted for additional confounders via inverse probability weights, we no longer observed an association between parental educational attainment and cognitive z-scores.

Respondent educational attainment was associated with domain-specific and global cognitive scores prior to adjustment for midlife poverty (Table 3, Model 2). Having more than a primary education but less than a high school education (vs. primary education or less) was associated with higher memory ({3: 0.33, 95% CI: 0.17, 0.50), executive function ({3: 0.44, 95% CI: 0.34, 0.54), verbal fluency ({3: 0.37, 95% CI: 0.22, 0.53), and global cognition z-scores ({3: 0.36, 95% CI: 0.25, 0.47). Estimates were of even larger magnitude when comparing those with a high school education, GED, or greater to those with a primary education or less (i.e., memory {3: 0.43, 95% CI: 0.23, 0.63; executive function {3: 0.67, 95% CI: 0.54, 0.79; verbal fluency {3: 0.69, 95% CI: 0.50, 0.88; global cognition {3: 0.58, 95% CI: 0.45, 0.71).

Estimates of association between greater than primary education (vs. primary education or less) and memory and verbal fluency z-scores were attenuated after including midlife poverty status in the model (Table 3, Model 3). Otherwise, controlled direct effect estimates for respondent education generally remained of similar magnitude after accounting for midlife poverty.

Associations between midlife poverty status and cognitive performance z-scores were modest to null; the lower bound of the 95% confidence interval crossed the null in all cases (i.e. memory {3: 0.08, 95% CI: -0.10, 0.25; executive function {3: 0.10, 95% CI: -0.02, 0.21; verbal fluency {3: -0.07, 95% CI: -0.24, 0.11; global cognition {3: 0.03, 95% CI: -0.09, 0.15).

All estimates were consistent in sensitivity analyses that excluded those who completed interviews by phone (Appendix Table 4), that excluded the Trails Making Test B in executive function and global cognitive z-scores (Appendix Table 5), and that modeled midlife poverty as a binary exposure, excluding those who had missing income data (Appendix Table 6). Estimates were also consistent in models that additionally included inverse probability of attrition weights to account for attrition from the study’s two baseline visits (1999/2000 for the original cohort, 2009 for the refresher cohort) (Appendix Table 7).

We next calculated the marginal difference in mean domain-specific and global cognitive scores for respondents in alignment with hypothetical lifecourse SES exposure scenarios (Figure 3; Appendix Table 8). Because of the relatively small sample sizes for contrasts that involved exposures across all three lifecourse periods (see Appendix Table 9) and because of our finding of no controlled direct estimate of association between early childhood SES and cognitive outcomes after accounting for childhood and midlife SES, we focused on scenarios that varied mid-childhood and midlife SES.

Relative SES disadvantage from mid-childhood to midlife (i.e., both primary education or less and midlife poverty) was associated with lower cognitive outcomes. Conversely, relative SES advantage from mid-childhood to midlife (vs. relative disadvantage across both periods) was associated with higher domain-specific and global cognitive z-scores (e.g. global cognition {3: 0.63, 95% CI: 0.49, 0.77). Those who experienced higher (> primary) education but had incomes at or below the FPL in midlife also had higher average domain-specific and global cognitive z-scores compared to those with both lower education and midlife poverty (e.g. global cognition {3: 0.51, 95% CI: 0.40, 0.61). The mean differences in domain-specific and global cognitive z-scores for those who experienced relatively low (i.e. ≤ primary) education but had household incomes above the federal poverty line were meaningfully different from those who experienced relative SES disadvantage across both measures. However, the magnitude of difference was much smaller as compared to the other contrasts across education and midlife SES patterns (e.g. global cognition {3: 0.12, 95% CI: 0.02, 0.23).

## DISCUSSION

Rigorous research on the relationship between lifecourse SES and cognitive aging that appropriately accounts for time-varying confounding has been limited to primarily non-Hispanic White, US-born samples with relatively high SES. We evaluated the relationship between SES measured at three points along the lifecourse (early-childhood, mid-childhood, and midlife) and midlife cognitive outcomes among a community-based study of primarily Latina, immigrant women living who have experienced relatively low lifecourse SES – a group historically excluded from studies of cognitive aging in the US.

We found mixed evidence in support of the pathways and sensitive periods lifecourse models, with less support for the accumulation model. Consistent with the pathways model, we found evidence of total associations but no controlled direct associations between parental educational attainment (our measure of early-childhood SES) and midlife cognitive outcomes after accounting for respondent education and midlife poverty. This is inconsistent with one of the few prior studies to apply marginal structural models in the context of lifecourse SES and cognitive aging, which found meaningful controlled direct effects of early-childhood SES.^5^ One of the notable differences between ours and prior studies is that our sample reported much lower parental (and own) educational attainment than the general US population, which may translate to fewer returns to intergenerational cognition. It could also be that the context of immigration renders the controlled direct effect estimates for *parental* educational attainment less meaningful for later-life cognitive outcomes given the structural disadvantages faced by immigrants in the labor market, which can often yield lateral or even downward occupational mobility relative to opportunities in Mexico.^31,32^

Consistent with the sensitive periods model, the magnitude of the direct association between respondent educational attainment and cognitive outcomes after accounting for midlife poverty was much larger than estimates corresponding other lifecourse periods. Conversely, being above (vs. at or below) the federal poverty line was only modestly associated with some cognitive outcomes, with confidence intervals that crossed the null in all cases. These findings are consistent with the two prior studies that used a similar approach in the context of lifecourse SES and cognitive aging,^2,5^ which also underscored the large magnitude of association for respondent educational attainment with cognitive performance vs. SES at other lifecourse stages. These findings may be due to several factors, including the occurrence of schooling at time of rapid brain development and the potential for education to instill abilities (e.g., literacy) that may yield lifelong opportunities for cognitive stimulation.^33^ In addition, there may not have been sufficient variability in household income among this sample, which experiences high rates of poverty and related structural disadvantages (e.g. low wages, high rates of food insecurity), to identify associations with midlife cognition.

We found substantial variation in mean cognitive scores given patterns of lifecourse SES exposure. Those who experienced relatively higher SES throughout the lifecourse had higher average cognitive scores relative to those who reported relatively low SES from mid-childhood to midlife. To place these estimates in context, we calculated that each year of additional age was associated with 0.018 SD decline in global cognition z-scores (95% CI: -0.030, -0.007). This means that compared to those who experienced relatively low lifecourse SES, those who experienced relatively high lifecourse SES had estimated (higher) cognitive scores equivalent to a difference in about 34 years of age. Contrasts between other sequences of lifecourse SES suggested that educational attainment appeared to be most important for driving differences in midlife cognitive scores.

### Limitations

First, we acknowledge that our study is based on a non-probability sample as part of a community-based study. By virtue of enrollment criteria for the original birth cohort, all participants were residing in Salinas Valley by the time of their index pregnancy, circa 1999-2001 and were low-income (e.g. Medicaid eligible). While participants resembled similarly aged immigrant women in the surrounding area, there were some notable differences, including higher rates of poverty (Appendix Table 2). Moreover, participants had been in the US for at least 20 years by the time of this study and more recent immigrants are not represented. Second, because participants were recruited from a longstanding cohort, there was attrition prior to the assessment focused on midlife cognition. Attrition from the study was to some extent differential with respect to lifecourse SES, although our estimates were robust to the inclusion of inverse probability of attrition weights.

Finally, we do not yet have repeated measures of cognitive performance that would enable the modeling of cognitive decline. Numerous studies have found that estimates of association between SES and *level* of cognitive performance often are not mirrored in estimates of association with the rate of cognitive decline.^34^ Understanding associations between lifecourse SES and age-related changes in cognitive performance among groups historically excluded from such research is of critical importance.

### Conclusion

In this community-based cohort of primarily immigrant Latina women, we estimated the relationship between lifecourse socio-economic status and midlife cognitive outcomes via marginal structural models. This work builds on prior studies^3,5^ that have applied similar methods to the study of lifecourse SES and cognitive outcomes, but that have been done in primarily White, US-born, and relatively high SES groups. Even among groups that have experienced relatively low lifecourse SES and related structural disadvantages, interventions to improve socio-economic status, even at the lower end of the SES distribution, can have a meaningful impact on brain health in midlife. Our results point to the importance of educational attainment for midlife brain health among a subgroup of individuals historically excluded from cognitive aging research.

## Data Availability

All data produced in the present study are available upon reasonable request to the authors, including completion of a data use agreement.

## Supplemental Appendix

**Appendix Table 1.** Comparison of Baseline CHAMACOS Sample Characteristics Overall and by CHAMACOS Maternal Cognition Study (MCS) participation.

|  | Baseline Sample<br>(1999 Cohort) (n<br>= 597) | Baseline<br>Sample<br>(2009<br>Cohort) (n =<br>635) | MCS<br>Sample<br>(n = 517) | Did Not<br>Participate<br>in MCS<br>(n = 315) |
| --- | --- | --- | --- | --- |
| **Educational attainment, n (%)** |  |  |  |  |
| 6th grade or less | 260 (43.6) | 272 (42.8) | 226 (43.7) | 153 (41.7) |
| Grades 7-12 | 218 (36.5) | 212 (33.4) | 176 (34.0) | 131 (35.7) |
| More than high school | 119 (19.9) | 151 (23.8) | 115 (22.2) | 83 (22.6) |
| **Nativity, n (%)** |  |  |  |  |
| US-born | 76 (12.7) | 78 (12.3) | 56 (10.9) | 52 (14.2) |
| Foreign born | 521 (87.1) | 557 (87.7) | 461 (89.2) | 315 (85.8) |
| **Race/ethnicity, n (%)** |  |  |  |  |
| Mexican, Mexican<br>American/Chicana, Mexican<br>Indian | 568 (94.9) | 600 (94.5) | 488 (94.4) | 341 (92.9) |
| Other Latina | 7 (1.2) | 9 (1.4) | 8 (1.5) | 6 (1.6) |
| Other<sup>b</sup> | 22 (3.7) | 22 (3.5) | 17 (3.3) | 20 (5.4) |
| Missing race/ethnicity | — | 4 (0.6) | 4 (0.8) | — |
| **Poverty status, n (%)** |  |  |  |  |
| At or below FPL | 368 (61.6) | 441 (69.4) | 362 (70.0) | 231 (62.9) |
| FPL-200% | 205 (34.3) | 182 (28.7) | 145 (28.0) | 122 (33.2) |
| >200% FPL | 22 (3.7) | 12 (1.9) | 10 (1.9) | 12 (3.3) |
| Missing income | 2 (0.3) | — | — | 2 (0.5) |
| **Marital status, n (%)** |  |  |  |  |
| Married/Living as married | 479 (80.2) | 518 (81.8) | 420 (81.2) | 288 (78.5) |
| Single | 86 (14.4) | 69 (10.9) | 62 (12.0) | 51 (13.9) |
| Separated | 26 (4.4) | 32 (5.1) | 23 (4.4) | 23 (6.3) |
| Divorced | 6 (1.0) | 12 (1.9) | 8 (1.5) | 5 (1.4) |
| Widowed | — | 2 (0.3) | 2 (0.4) | — |
| Missing | — | 3 (0.5) | 2 (0.4) | — |
| **Employment status, n (%)** |  |  |  |  |
| Worked since last visit<br>(agriculture) | 65 (10.9) | 61 (9.7) | 49 (9.5) | 38 (10.4) |
| Worked since last visit (non-<br>agriculture) | 269 (45.1) | 307 (48.3) | 250 (48.4) | 168 (45.8) |
| Did not work since last visit | 262 (43.9) | 264 (41.8) | 215 (41.6) | 160 (43.6) |
| Missing | 1 (0.2) | 3 (0.4) | 3 (0.6) | 1 (0.3) |
| **Composite health score, n (%)<sup>a</sup>** |  |  |  |  |
| None | 516 (86.4) | 503 (79.2) | 405 (78.3) | 313 (85.3) |
| 1+ | 81 (13.6) | 132 (20.8) | 112 (21.7) | 54 (14.7) |
| Note: a. |  |  |  |  |

**Appendix Table 2.** Comparison of CHAMACOS Maternal Cognition Study participants to similarly-aged women in the Salinas Valley.

|  | Maternal Cognition<br>Study Participants<br>(n = 517) | Population<br>estimate<br>(n = 47,392)<sup>a</sup> | Population estimate,<br>foreign- born only<br>(n = 26,219)<sup>a</sup> |
| --- | --- | --- | --- |
| **Educational attainment, n (%)** |  |  |  |
| 6th grade or less | 226 (43.7) | 11,501 (24.3) | 11,148 (42.5) |
| Grades 7-12 | 176 (34.0) | 16,455 (34.7) | 9,415 (35.9) |
| More than high school | 115 (22.2) | 19,436 (41.0) | 5,656 (21.6) |
| **Nativity, n (%)** |  |  |  |
| US-born | 56 (10.9) | 21,173 (44.7) | — |
| Foreign-born | 459 (89.1) | 26,219 (55.3) | — |
| Missing country of birth | 2 (0.4) | — | — |
| **Race and ethnicity, n (%)** |  |  |  |
| Mexican/Mexican-American | 488 (94.4) | 30,579 (64.5) | 21,927 (83.6) |
| Other Latina | 8 (1.5) | 2,130 (4.5) | 1,499 (5.7) |
| Other<sup>b</sup> | 17 (3.3) | 14,683 (31.0) | 2,793 (10.7) |
| Missing race and ethnicity | 4 (0.8) | — | — |
| **Language spoken at home, n (%)<sup>c</sup>** |  |  |  |
| Spanish | 482 (93.2) | 30,776 (64.9) | 23,325 (89.0) |
| English | 32 (6.2) | 13,553 (28.6) | 756 (2.9) |
| Other | 3 (0.6) | 3,063 (6.5) | 2,138 (8.2) |
| **Poverty category, n (%)<sup>d</sup>** |  |  |  |
| At or below FPL | 183 (38.2) | 5,105 (10.8) | 3,098 (11.8) |
| FPL-200% | 270 (56.4) | 7,983 (16.8) | 6,037 (23.0) |
| >200% FPL | 26 (5.4) | 34,304 (72.4) | 17,084 (65.2) |
| Missing income | 38 (7.4) | — | — |
| **Marital status, n (%)<sup>d</sup>** |  |  |  |
| Married/Living as married | 365 (70.6) | 29,919 (63.1) | 17,431 (66.5) |
| Single | 74 (14.3) | 8,868 (18.7) | 5,107 (19.5) |
| Separated | 37 (7.2) | 1,459 (3.1) | 778 (3.0) |
| Divorced | 35 (6.8) | 5,991 (12.6) | 2,215 (8.5) |
| Widowed | 6 (1.2) | 1,155 (2.4) | 688 (2.6) |
| **Employment status, n (%)<sup>d</sup>** |  |  |  |
| Worked since last visit (agriculture) | 247 (47.8) | 6,559 (13.8) | 6,411 (24.5) |
| Worked since last visit (non-agricultural) | 165 (31.9) | 30,713 (64.8) | 13,507 (51.5) |
| Did not work since last visit | 105 (20.3) | 10,119 (21.4) | 6,301 (24.0) |
| **People per household, n (%)<sup>d</sup>** |  |  |  |
| 0-5 | 352 (68.1) | 37,024 (78.1) | 18,724 (71.4) |
| 6+ | 164 (31.7) | 10,368 (21.9) | 7,495 (28.6) |
| Missing | 1 (0.2) | — | — |
| Notes: a. From 2021 American Community Survey 5-year Public Use Microsample (PUMs) Data. Geographies used included Monterey County Northeast (Salinas City) and Monterey County East+South + San Benito County; b. Other includes Non-Hispanic White, Non-Hispanic Black, Asian/Pacific Islander, and Other; c. n=16 participants stated they spoke English and Spanish equally at baseline and n=2 were missing language spoken at baseline. These participants are classified by the language of their Maternal Cognition interview (n=14 English and n=4 Spanish); d. Data on time-varying characteristics of the CHAMACOS Maternal Cognition Study was collected between 2022-2023. |  |  |  |

**Appendix Table 3.**
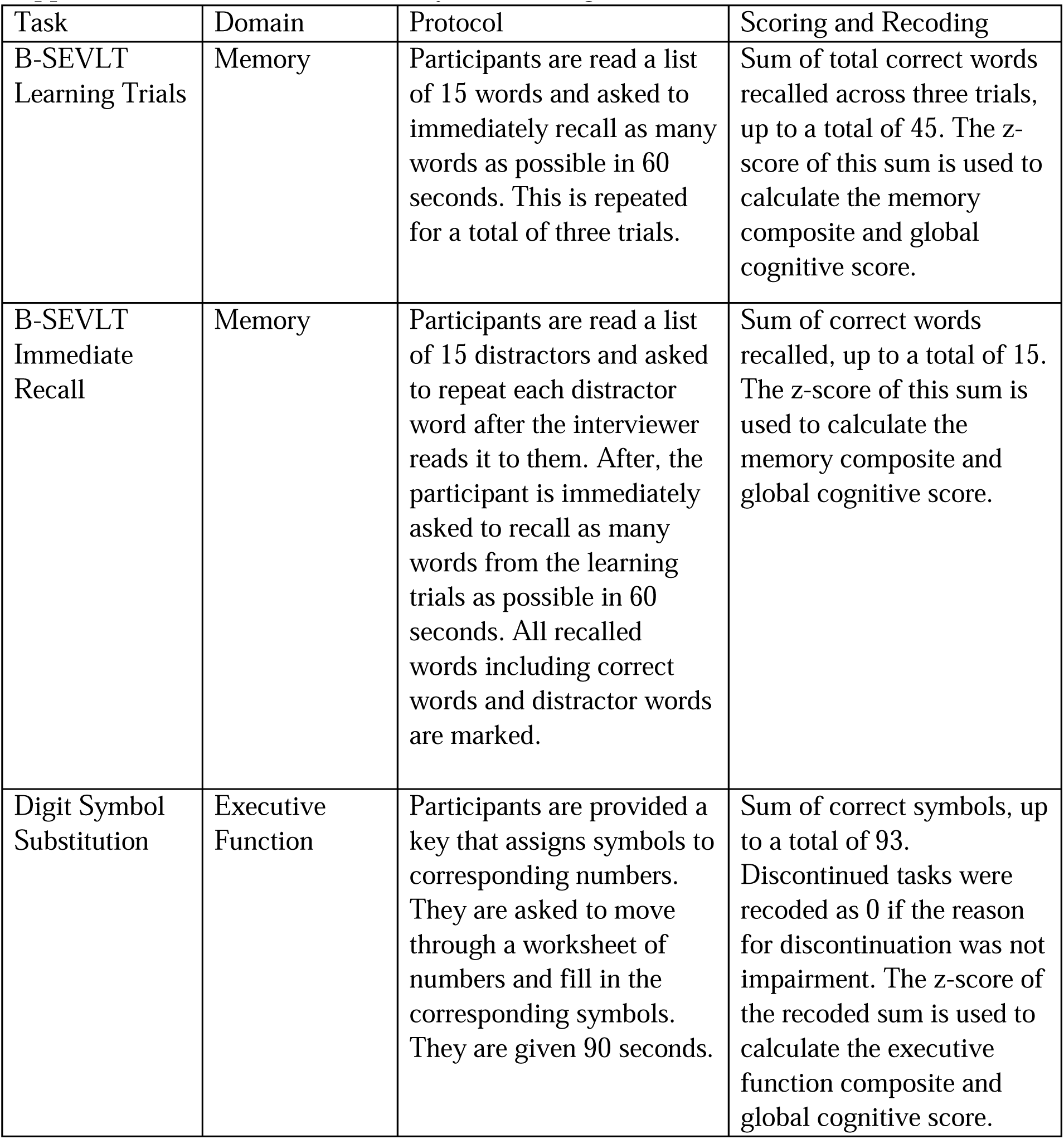
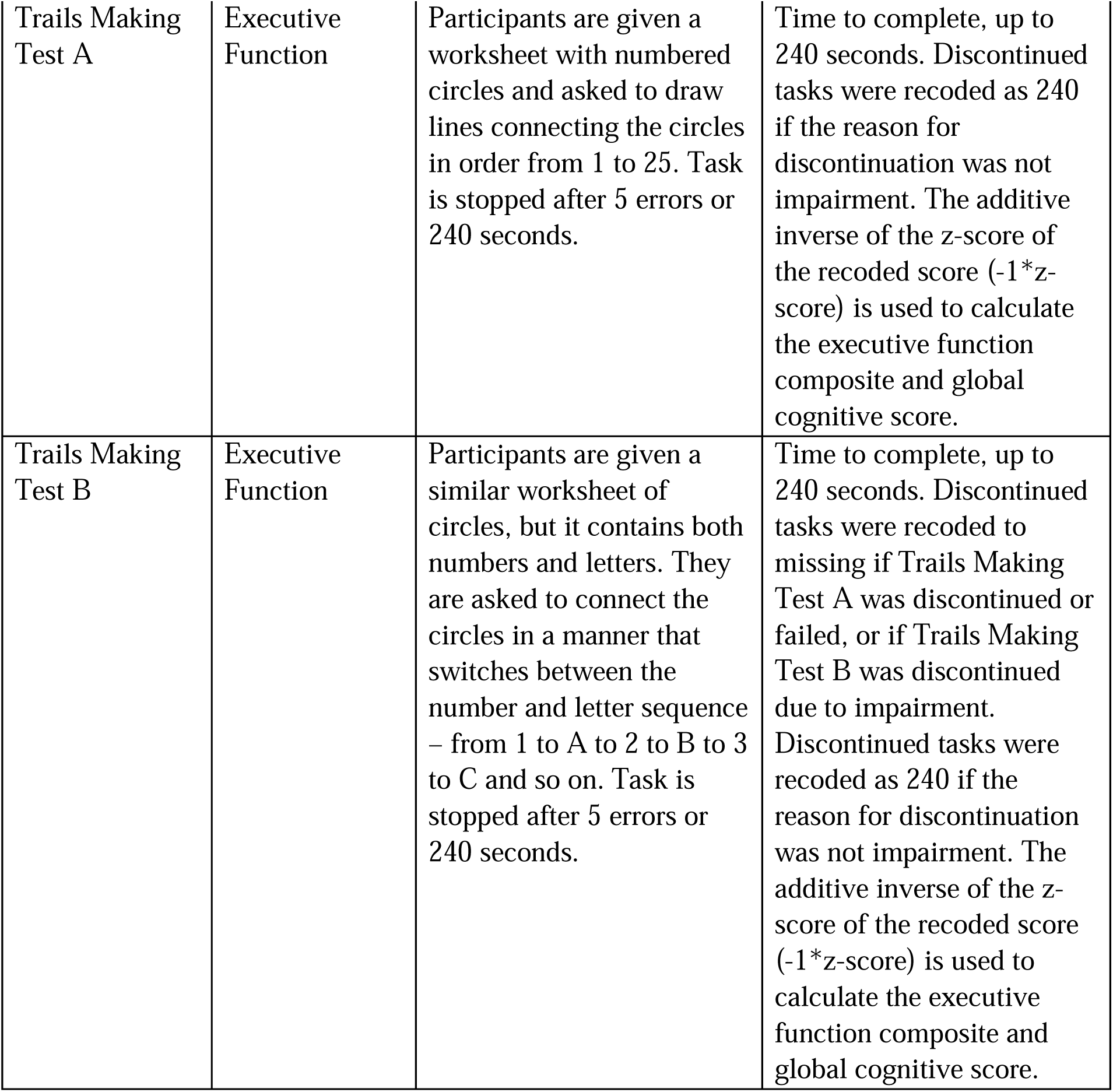
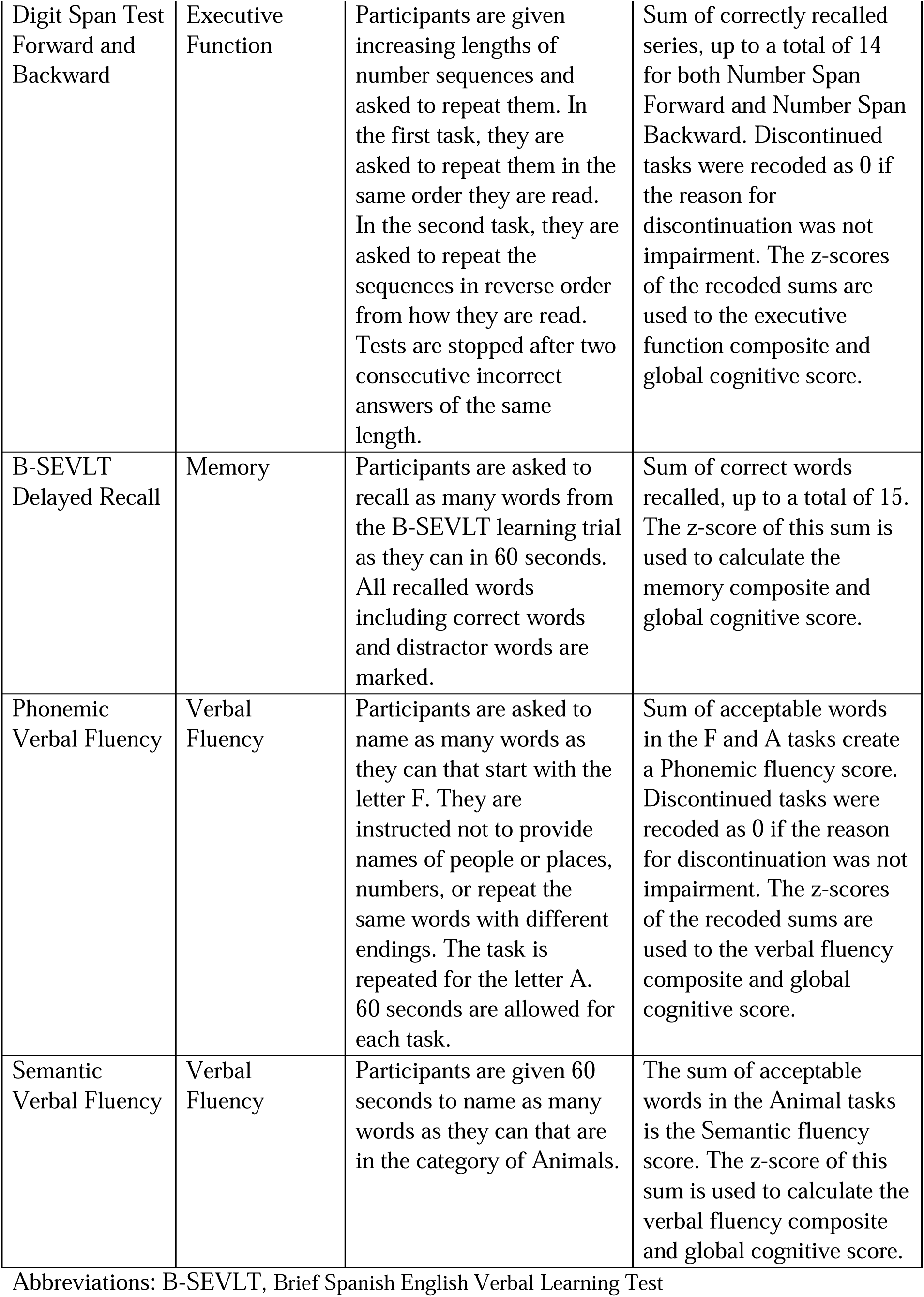
Detailed Summary of Neurocognitive Tasks.

**Appendix Table 4.**
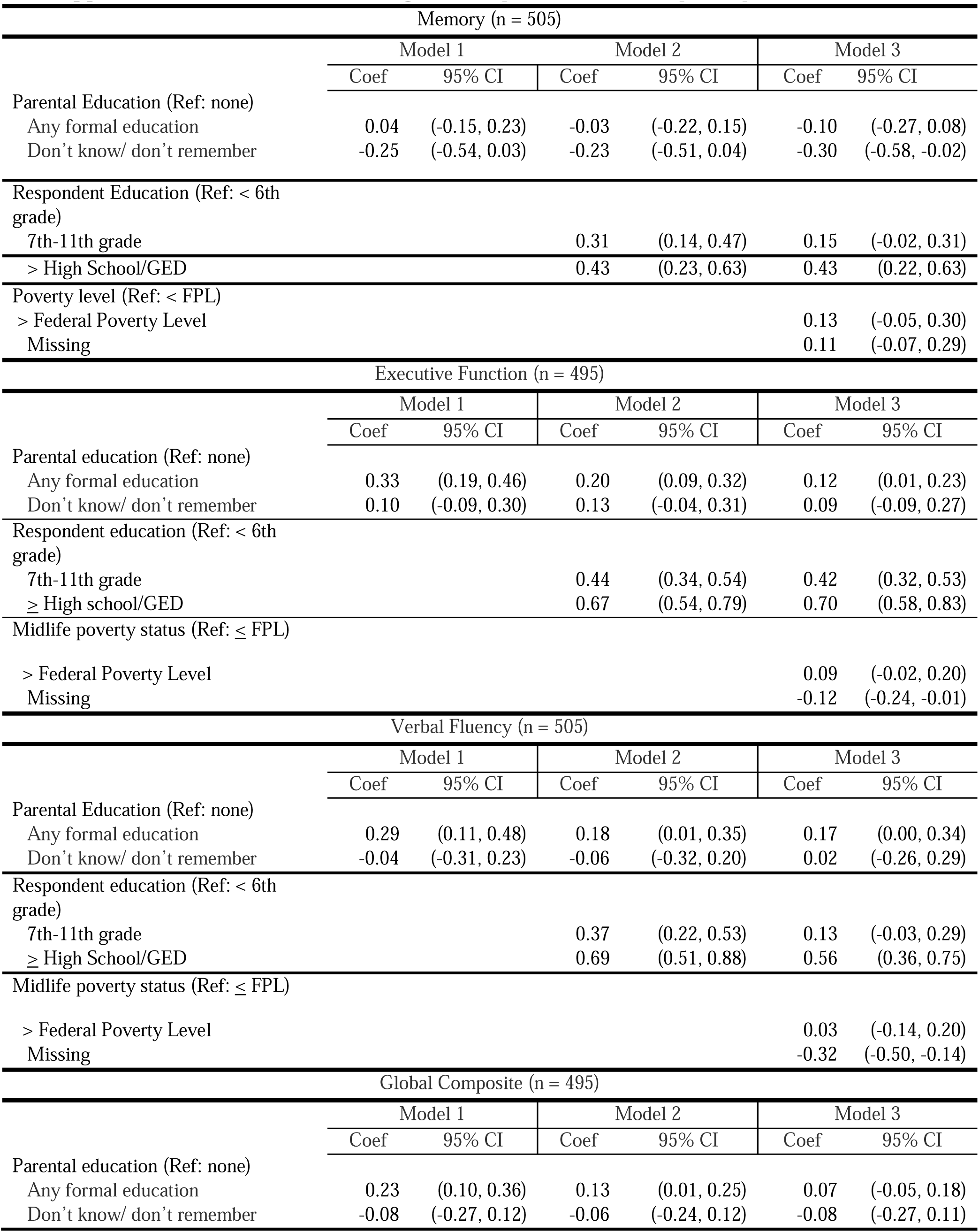
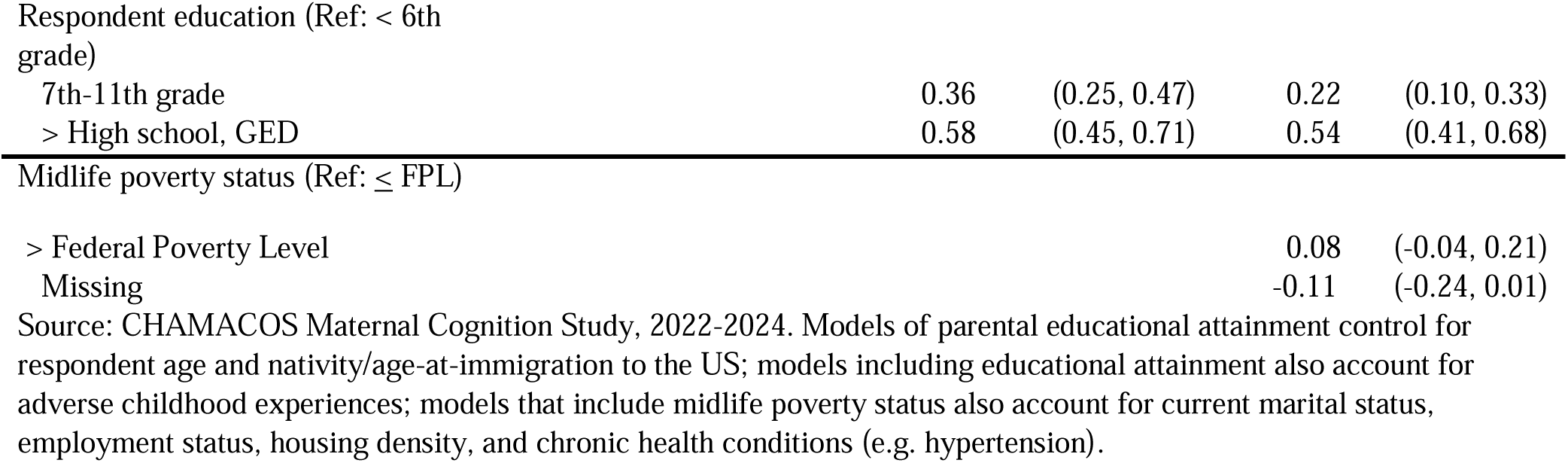
Results excluding n = 6 respondents who completed phone visits.

**Appendix Table 5.**
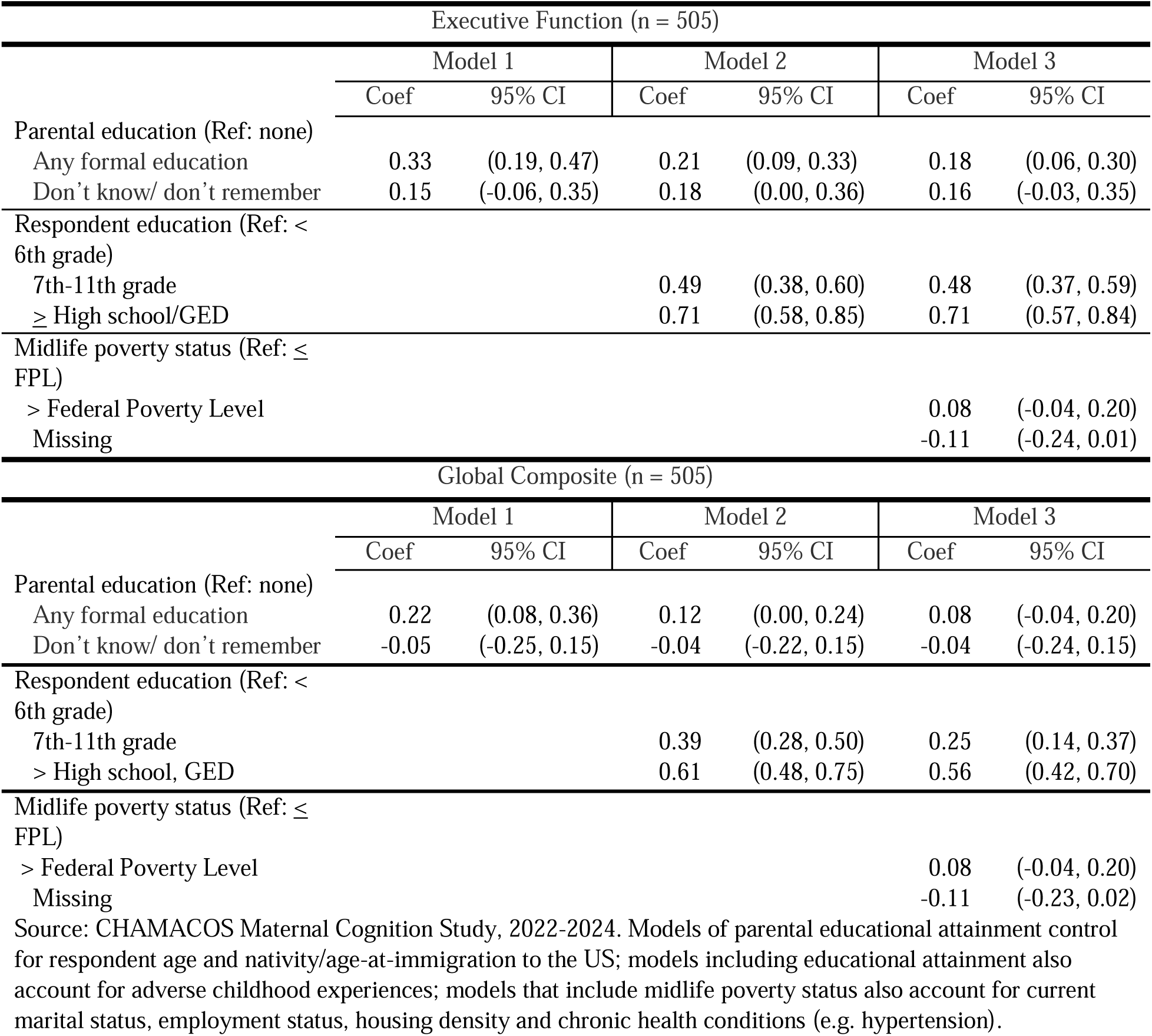
Results excluding Trails Making Test B from executive function and global composite scores.

| Executive Function (n = 505) |  |  |  |  |  |  |
| --- | --- | --- | --- | --- | --- | --- |
|  | Model 1 |  | Model 2 |  | Model 3 |  |
|  | Coef | 95% CI | Coef | 95% CI | Coef | 95% CI |
| Parental education (Ref: none) |  |  |  |  |  |  |
| Any formal education | 0.33 | (0.19, 0.47) | 0.21 | (0.09, 0.33) | 0.18 | (0.06, 0.30) |
| Don't know/ don't remember | 0.15 | (-0.06, 0.35) | 0.18 | (0.00, 0.36) | 0.16 | (-0.03, 0.35) |
| Respondent education (Ref: < 6th grade) |  |  |  |  |  |  |
| 7th-11th grade |  |  | 0.49 | (0.38, 0.60) | 0.48 | (0.37, 0.59) |
| ≥ High school/GED |  |  | 0.71 | (0.58, 0.85) | 0.71 | (0.57, 0.84) |
| Midlife poverty status (Ref: ≤ FPL) |  |  |  |  |  |  |
| > Federal Poverty Level |  |  |  |  | 0.08 | (-0.04, 0.20) |
| Missing |  |  |  |  | -0.11 | (-0.24, 0.01) |
| Global Composite (n = 505) |  |  |  |  |  |  |
|  | Model 1 |  | Model 2 |  | Model 3 |  |
|  | Coef | 95% CI | Coef | 95% CI | Coef | 95% CI |
| Parental education (Ref: none) |  |  |  |  |  |  |
| Any formal education | 0.22 | (0.08, 0.36) | 0.12 | (0.00, 0.24) | 0.08 | (-0.04, 0.20) |
| Don't know/ don't remember | -0.05 | (-0.25, 0.15) | -0.04 | (-0.22, 0.15) | -0.04 | (-0.24, 0.15) |
| Respondent education (Ref: < 6th grade) |  |  |  |  |  |  |
| 7th-11th grade |  |  | 0.39 | (0.28, 0.50) | 0.25 | (0.14, 0.37) |
| > High school, GED |  |  | 0.61 | (0.48, 0.75) | 0.56 | (0.42, 0.70) |
| Midlife poverty status (Ref: ≤ FPL) |  |  |  |  |  |  |
| > Federal Poverty Level |  |  |  |  | 0.08 | (-0.04, 0.20) |
| Missing |  |  |  |  | -0.11 | (-0.23, 0.02) |

Source: CHAMACOS Maternal Cognition Study, 2022-2024. Models of parental educational attainment control for respondent age and nativity/age-at-immigration to the US; models including educational attainment also account for adverse childhood experiences; models that include midlife poverty status also account for current marital status, employment status, housing density and chronic health conditions (e.g. hypertension).

**Appendix Table 6.**
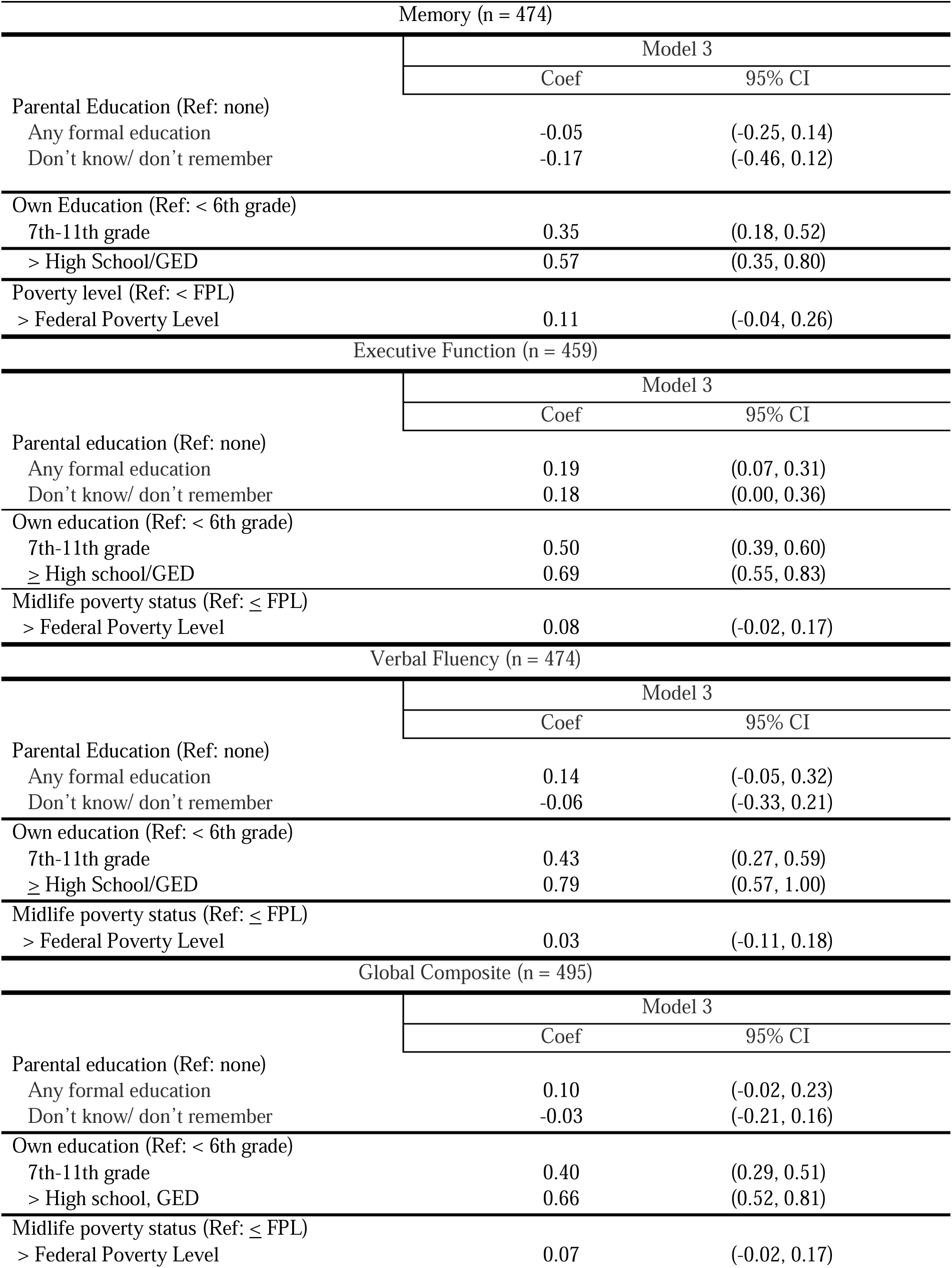
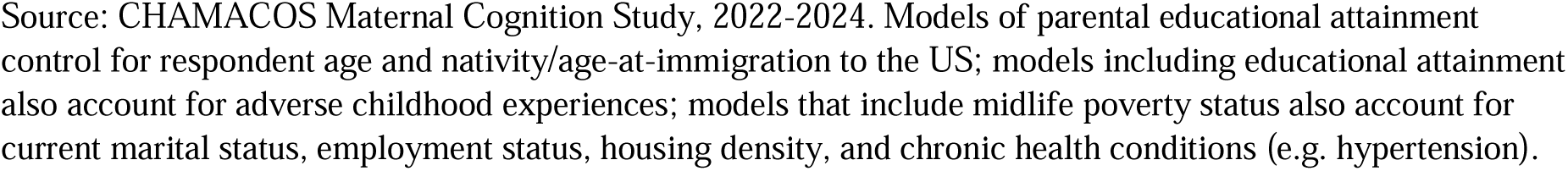
Results with Binary Poverty Measure, Removing Missing Values.

| Memory (n = 474) |  |  |
| --- | --- | --- |
|  | Model 3 |  |
|  | Coef | 95% CI |
| Parental Education (Ref: none) |  |  |
| Any formal education | -0.05 | (-0.25, 0.14) |
| Don't know/ don't remember | -0.17 | (-0.46, 0.12) |
| Own Education (Ref: < 6th grade) |  |  |
| 7th-11th grade | 0.35 | (0.18, 0.52) |
| > High School/GED | 0.57 | (0.35, 0.80) |
| Poverty level (Ref: < FPL) |  |  |
| > Federal Poverty Level | 0.11 | (-0.04, 0.26) |
| Executive Function (n = 459) |  |  |
|  | Model 3 |  |
|  | Coef | 95% CI |
| Parental education (Ref: none) |  |  |
| Any formal education | 0.19 | (0.07, 0.31) |
| Don't know/ don't remember | 0.18 | (0.00, 0.36) |
| Own education (Ref: < 6th grade) |  |  |
| 7th-11th grade | 0.50 | (0.39, 0.60) |
| ≥ High school/GED | 0.69 | (0.55, 0.83) |
| Midlife poverty status (Ref: ≤ FPL) |  |  |
| > Federal Poverty Level | 0.08 | (-0.02, 0.17) |
| Verbal Fluency (n = 474) |  |  |
|  | Model 3 |  |
|  | Coef | 95% CI |
| Parental Education (Ref: none) |  |  |
| Any formal education | 0.14 | (-0.05, 0.32) |
| Don't know/ don't remember | -0.06 | (-0.33, 0.21) |
| Own education (Ref: < 6th grade) |  |  |
| 7th-11th grade | 0.43 | (0.27, 0.59) |
| ≥ High School/GED | 0.79 | (0.57, 1.00) |
| Midlife poverty status (Ref: ≤ FPL) |  |  |
| > Federal Poverty Level | 0.03 | (-0.11, 0.18) |
| Global Composite (n = 495) |  |  |
|  | Model 3 |  |
|  | Coef | 95% CI |
| Parental education (Ref: none) |  |  |
| Any formal education | 0.10 | (-0.02, 0.23) |
| Don't know/ don't remember | -0.03 | (-0.21, 0.16) |
| Own education (Ref: < 6th grade) |  |  |
| 7th-11th grade | 0.40 | (0.29, 0.51) |
| > High school, GED | 0.66 | (0.52, 0.81) |
| Midlife poverty status (Ref: ≤ FPL) |  |  |
| > Federal Poverty Level | 0.07 | (-0.02, 0.17) |

Source: CHAMACOS Maternal Cognition Study, 2022-2024. Models of parental educational attainment control for respondent age and nativity/age-at-immigration to the US; models including educational attainment also account for adverse childhood experiences; models that include midlife poverty status also account for current marital status, employment status, housing density, and chronic health conditions (e.g. hypertension).

**Appendix Table 7.**
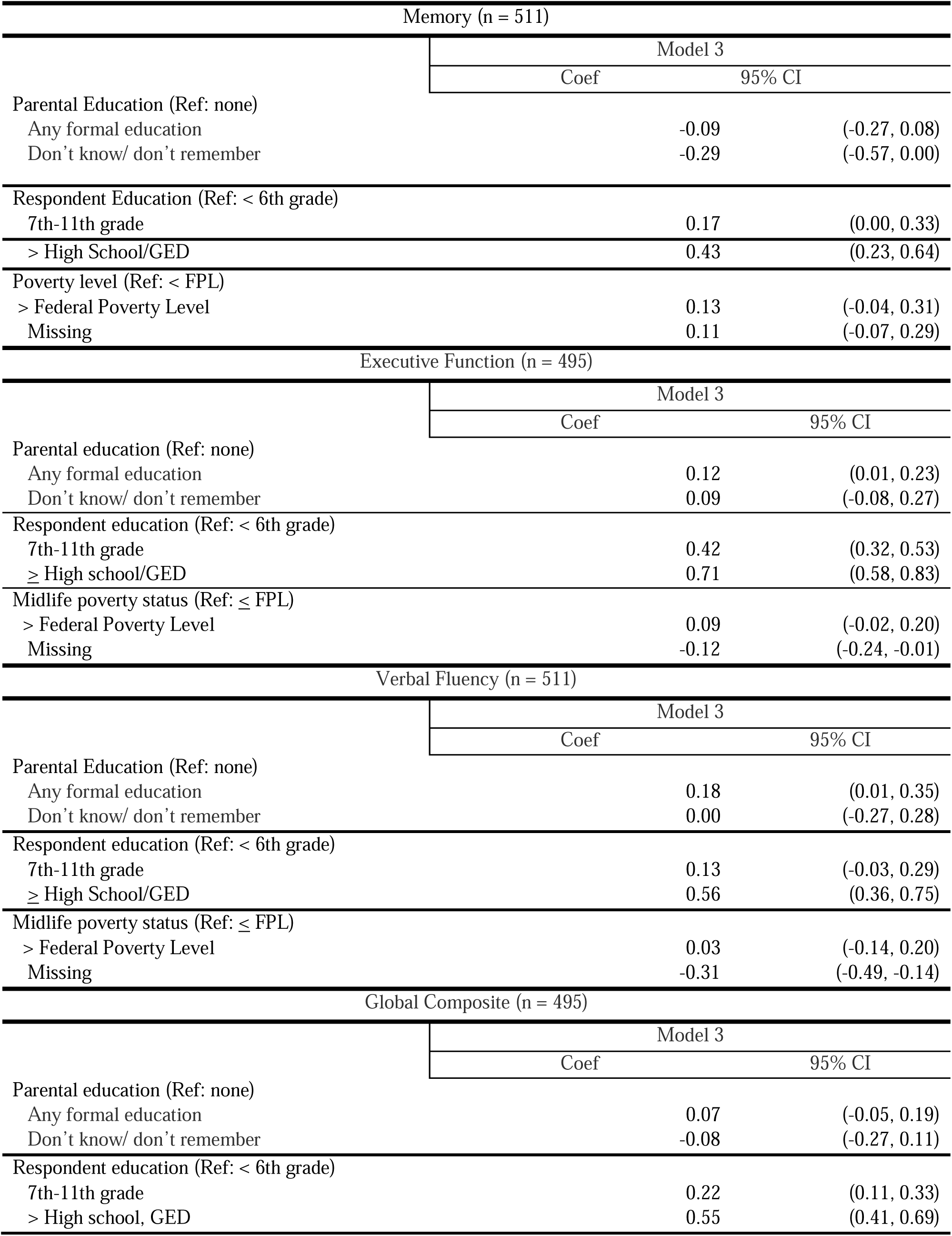
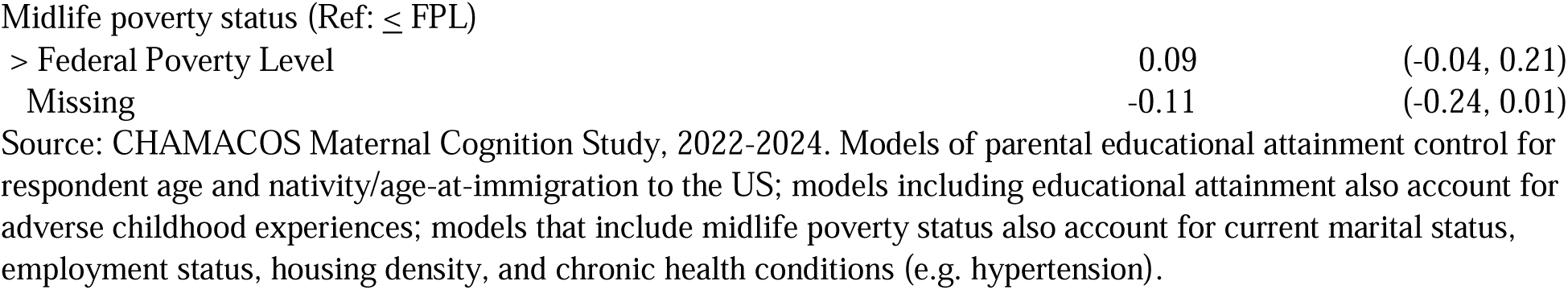
Table of Results with Inverse Probability of Attrition Weights.

**Appendix Table 8.** Mean difference in each cognitive z-score between stable low respondent education and poverty status (<=6^th^ grade education and at or below poverty level) and all combinations of respondent education and poverty status.

| **Outcome** | **Respondent Education** | **Current Poverty Level** | **Beta** | **95% CI** |
| --- | --- | --- | --- | --- |
| Memory Composite | $\leq 6^{\text{th}}$ grade | Above poverty level | 0.131 | (-0.030, 0.292) |
| Memory Composite | $\geq 7^{\text{th}}$ grade | At or below poverty level | 0.422 | (0.254, 0.590) |
| Memory Composite | $\geq 7^{\text{th}}$ grade | Above poverty level | 0.553 | (0.339, 0.767) |
| Executive Function | $\leq 6^{\text{th}}$ grade | Above poverty level | 0.118 | (0.014, 0.221) |
| Executive Function | $\geq 7^{\text{th}}$ grade | At or below poverty level | 0.588 | (0.489, 0.688) |
| Executive Function | $\geq 7^{\text{th}}$ grade | Above poverty level | 0.706 | (0.564, 0.847) |
| Verbal Fluency | $\leq 6^{\text{th}}$ grade | Above poverty level | 0.127 | (-0.023, 0.276) |
| Verbal Fluency | $\geq 7^{\text{th}}$ grade | At or below poverty level | 0.558 | (0.409, 0.707) |
| Verbal Fluency | $\geq 7^{\text{th}}$ grade | Above poverty level | 0.685 | (0.483, 0.887) |
| Global Function | $\leq 6^{\text{th}}$ grade | Above poverty level | 0.124 | (0.017, 0.231) |
| Global Function | $\geq 7^{\text{th}}$ grade | At or below poverty level | 0.505 | (0.400, 0.611) |
| Global Function | $\geq 7^{\text{th}}$ grade | Above poverty level | 0.629 | (0.487, 0.772) |

**Appendix Table 9.** Table of Lifecourse SES Exposure Patterns for CHAMACOS Maternal Cognition Study Participants.

|  | Overall<br>(n = 511) |  | Parents completed no<br>formal education or<br>education unknown<br>(n = 175) |  | Parents completed<br>any formal<br>education (n = 336) |  |
| --- | --- | --- | --- | --- | --- | --- |
| Respondent Education +<br>Current Poverty Levels | n | % | n | % | n | % |
| Primary or below + at or<br>below poverty level | 90 | 17.6 | 45 | 25.7 | 45 | 13.4 |
| Primary or below + above<br>poverty level | 111 | 21.7 | 46 | 26.3 | 65 | 19.3 |
| Above primary + at or<br>below poverty level | 92 | 18.0 | 24 | 13.7 | 68 | 20.2 |
| Above primary + above<br>poverty level | 181 | 35.4 | 41 | 23.4 | 140 | 41.7 |
| Missing (driven by<br>missing on poverty) | 37 | 7.2 | 19 | 10.9 | 18 | 5.4 |

### Appendix Text 1. Hypertension Classification

We classified individuals as having hypertension if they self-reported a prior doctor diagnosis or were classified as having a systolic blood pressure of ≥ 120mm/Hg or a diastolic blood pressure ≥ 80 mm/Hg based on in-office blood pressure measurements. We measured resting blood pressure at three 1-minute intervals using an automatic digital sphygmomanometer (Dianamap Carescape V100, Avante Health Solutions, Chicago IL); the last two measurements were averaged for analysis. Individuals who had originally self-reported no prior doctor diagnosis of hypertension were re-classified as having hypertension from their blood pressure measurements.

### Appendix Text 2. Description of Diabetes Classification

To classify individuals as ever being diabetic, we used a combination of self-reported diabetes information as well as fasting glucose measures and/or HBA1c scores. Individuals who self-reported diabetes, those who had fasting glucose levels higher than 125 mg/dL, and those who had an HBA1c score greater than 6.5% (among those who did not have a fasting blood glucose lab) were classified as having diabetes.

### Appendix Text 3. Calculating inverse probability weights

The first exposure model estimated the probability of the participant having above a primary school education (vs. primary school or less) conditional on age, nativity/age-at-migration, language of assessment, parental educational attainment, and a continuous measure of adverse childhood experiences. The model took the following form:

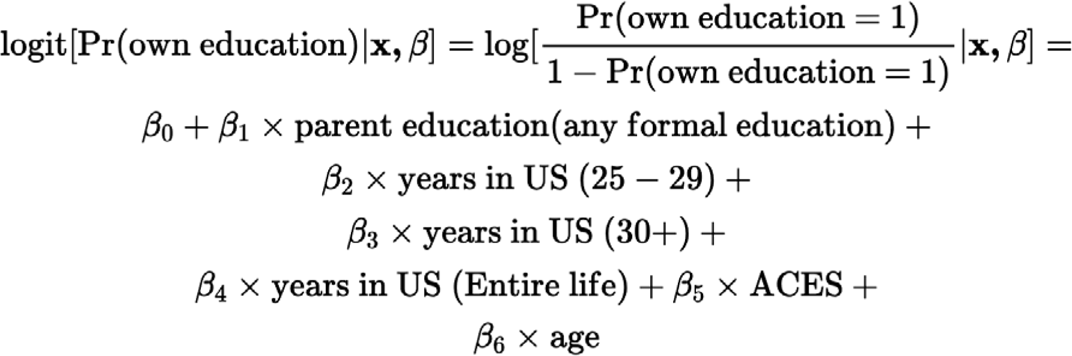

Where the reference level for parental education was no formal education and the reference level for years in the US was 20-24 years. We compared the model above to an expanded model that included interaction terms between the exposure of interest (parental education) and all covariates. A comparison of the two models via an ANOVA likelihood ratio test yielded a p-value of 0.11. We therefore proceeded to use the original simplified model that did not contain any interaction variables.

We proceeded with the exposure model that estimated the probability of the participant reporting a household income above the poverty level relative to at or below the poverty level and the probability of missing household income relative to income at or below the poverty level using multinomial logistic regression. We compared this model to a model that included interaction terms using a likelihood ratio test; based on those results, we proceeded to use the original simplified model that did not contain any interaction variables. We calculated the overall weights as the product of the weights from the education exposure model and the weights from the poverty exposure model. We trimmed the combined weights at the 99^th^ percentile.

### Appendix Text 4. Calculating inverse probability of attrition weights

We used inverse probability of attrition weights to up-weight the characteristics of those who were lost to follow-up and down-weight the characteristics of those who were retained throughout the study. We calculated these weights as the probability the individual participated in the maternal cognition study as a function of the mom’s age at the maternal cognition study initiation, the age the mom came to the US, the mom’s educational attainment, and baseline variables including marital status, work status as pregnancy/baseline, poverty status, and composite health score. We also included interactions between the mother’s cohort and time-varying variables (marital status, work status, poverty status, and composite health score). We multiplied these inverse probability of attrition weights by the original combined inverse probability of treatment weights to calculate an overall weight.

**Appendix Figure 1.**
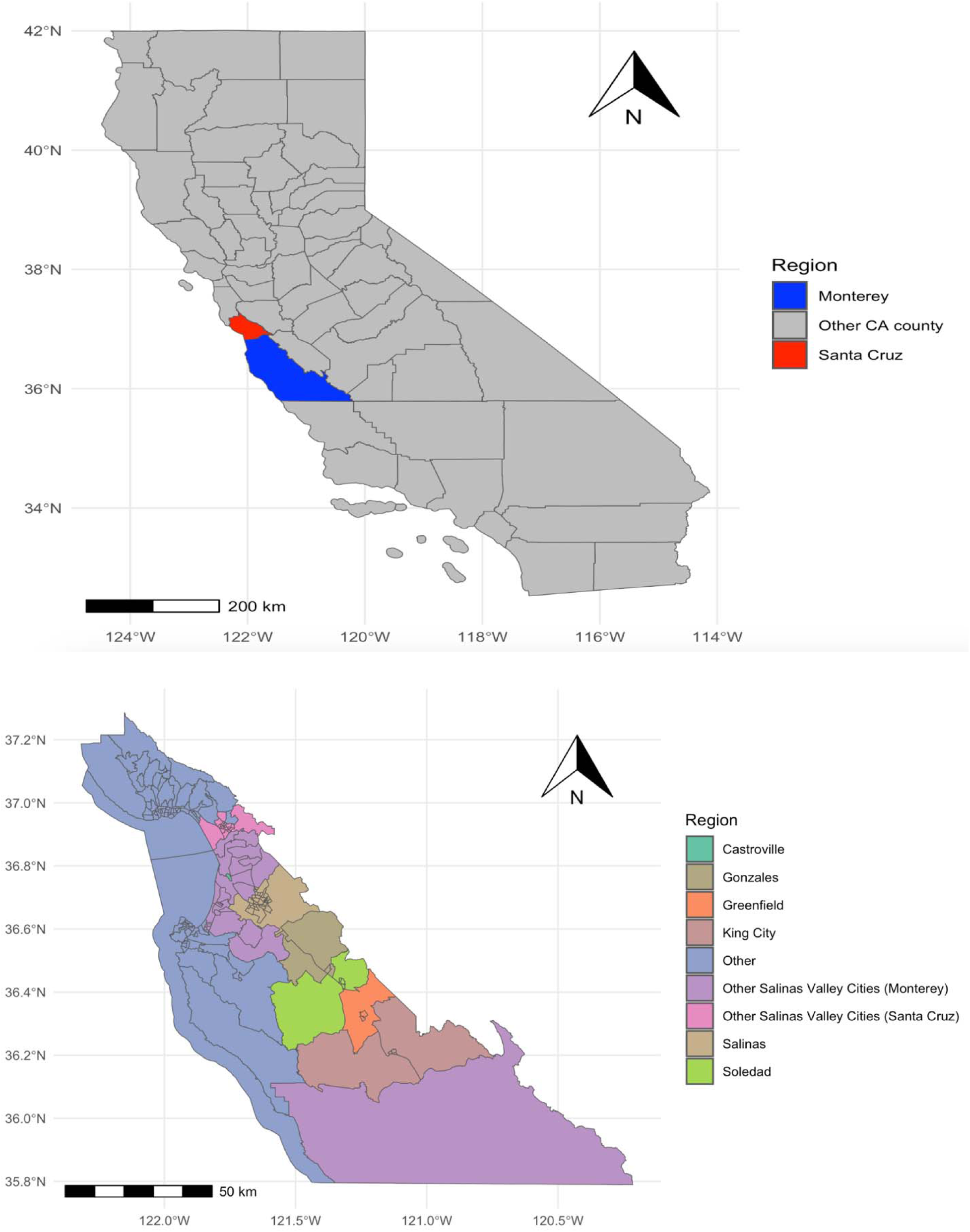
Map.

**Appendix Figure 2.**
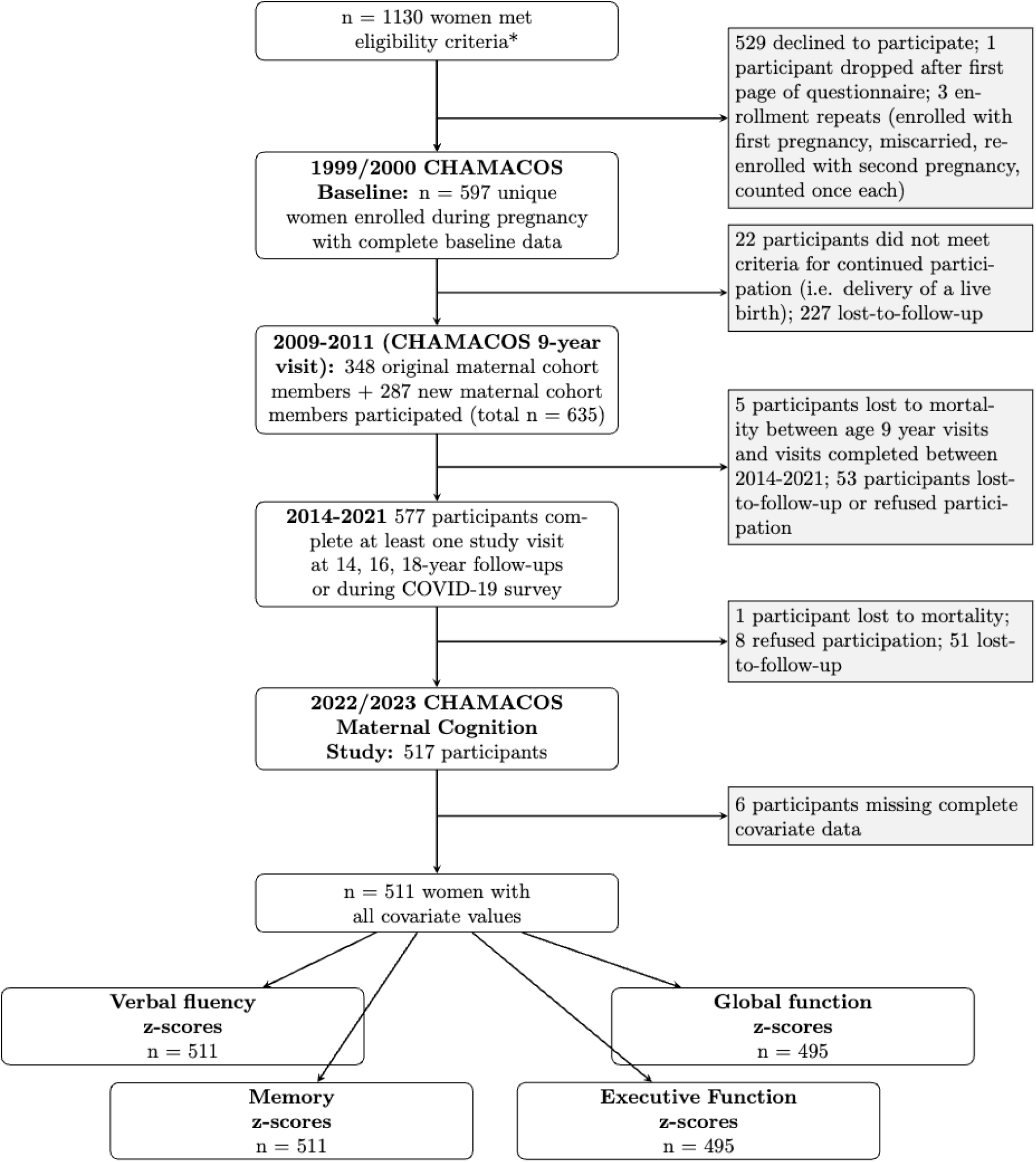
Study Flow Chart.

